# Near point of care C-Reactive Protein and Serum Amyloid-A Test met WHO target product profile for TB screening

**DOI:** 10.64898/2026.11.10.26367691

**Authors:** Nadya Iswandari, Todia Pediatama Setiabudiawan, Marcellus Korompis, Raspati C Koesoemadinata, Evan Susandi, Iceu D Kulsum, Dewi K Turbawaty, Bachti Alisjahbana

## Abstract

**Background:** TB eradication strategies in high burden countries need affordable, scalable, and accessible screening tools. This study investigated the diagnostic accuracy of C-reactive protein (CRP), serum amyloid A (SAA), and interferon gamma-induced protein 10 (IP-10) as potential screening tools.

**Method:** This cross-sectional diagnostic study investigated stored archived serum samples from three groups of subjects; bacteriology confirmed symptomatic TB group (n=57), probable TB (n=56) and healthy controls (n=60). CRP and SAA concentrations were measured using near point-of-care fluorescence immunoassay (NPOCT-FIA), while IP-10 was measured using enzyme-linked immunosorbent assay (ELISA). Diagnostic performance of individual and combined biomarkers was evaluated

**Results:** Receiver operating characteristic analysis demonstrated excellent diagnostic performance of serum amyloid A (SAA) and C-reactive protein (CRP) for identifying confirmed tuberculosis (TB). Compared with probable TB and healthy controls, SAA achieved an AUC of 0.961 (cut-off *>*31.0 mg/L), with 96.5% sensitivity and 88.6% specificity, while CRP achieved an AUC of 0.953 (cut-off 21.3 mg/L), with 96.5% sensitivity and 86.0% specificity. Interferon gamma-induced protein 10 (IP-10) showed poor discrimination (AUC 0.545). Combined CRP and SAA achieved 100.0% sensitivity and 83.3% specificity.

**Conclusion:** SAA and CRP met WHO target product profile criteria for TB triage screening.

## 1. Introduction

Tuberculosis remains a major global public health burden, with Indonesia accounting for the second-largest number of TB cases worldwide. Early detection through screening is essential to interrupt transmission and improve treatment out-comes. The World Health Organization (WHO) recommends population-based TB screening in settings with a prevalence of 0.5%, including Indonesia.(1) Indonesia is supposed to detect and treat one million TB cases annually to support TB elimination by 2030, but so far, only 81% of the target has been achieved. To address this substantial gap, the Indonesian Ministry of Health has expanded active and intensified screening efforts in the community. Some researchers even recommended mass screening nationwide.(2,3,4)

Current screening strategies rely on clinical symptom assessment however, up to 10–25% of patients may be asymptomatic. Chest X-ray is recommended but often inaccessible in primary care especially resource-limited or remote settings.(4) Using sputum sample for screening is hard to attain due to the fact that 20% patients did not experience a productive cough or were unable to produce sputum. Although Nucleic Acid Amplification test (NAAT) based NPOCT are becoming more available with increasing accuracy and usability, it is still relatively expensive for mass screening. And there are a substantial proportion of symptomatic and extrapulmonary TB patient with lacking bacterial confirmation. Antigen Mtb, urine LAM and Mtb-Ag 85 and volatile organic compound test did not achieve desirable accuracy.(5,6,7) Screening tools which are preferably easier to use and affordable in a point-of-care test are needed for triage in community use.

Inflammatory host response against tuberculosis has been quite extensively studied. Several biomarkers were stated as potential e.g. CRP, Neural Cell Adhesion Molecule (NCAM), Serum Amyloid P (SAP), IP-10 (also known as CXCL10), Ferritin, tissue plasminogen activator (TPA), I-309 and monokine induced by gamma interferon (MIG).(8)

Our interest comes to two of the most known Biomarker which is CRP and SAA which are known to have near point of care test available commercially.

CRP is an acute inflammatory protein which increased and decrease proportionally to Tuberculosis disease progression. CRP is recommended by WHO for TB screening in HIV patients, (9,10,11,12) but not in non-HIV TB patients due to low specificity. Several other conditions associated with heightened CRP levels, such as other bacterial infections, autoimmune and malignancy.(13) Several studies concluded CRP have adequate sensitivity but the low specificity is inhibiting its wider use. Point-of-care test for CRP is commercially available as a fluorescent immunoassay platform as well as a qualitative lateral flow rapid test device.

SAA is an inflammatory apolipoprotein which can show increase in response to infection, trauma or malignancy. In infectious disease SAA has been utilized for screening of existing bloodstream infection.(14) But studies evaluating the capability of SAA to detect Tuberculosis are lacking. However, one study in Brazil showed promising sensitivity and specificity result of SAA differentiating patients with pulmonary TB and negative bacilloscopy group.(15,16) Since the test is already developed and commercially distributed as a NPOCT test, we include SAA in our evaluation.

With the intention to increase specificity, we include IP-10.8 in our study due to the possible linked with IFN-(interferon gamma) release in TB infection. IP-10 is a chemokine by product of IFN-which is secreted by T lymphocyte cells as a response to Tuberculosis infection (17) .IP-10 test shows promising specificity despite lower sensitivity.18 Combination of these biomarkers may be the solution to achieve target product profile accuracy by World Health Organization.

This study aimed to evaluate the diagnostic accuracy of NPOCT CRP, SAA, IP-10 ELISA and their combination as potential screening biomarkers for tuberculosis.

## 2. Methods

### Study Design

This cross-sectional diagnostic study investigates bio archive serum from the EVIDENT (EvaluatIon and DemonstratioN of new TB diagnostics for Indonesia) study conducted in June 2024 and June 2025 and a Covid-19 Vaccine study subjects conducted in Bandung, Indonesia, between 2021-2022.

### Study population

Between February and August 2018, we enrolled clients aged ≥ 15 years registering for any type of health services at the PHC, proportional to each PHC’s estimated catchment population. We excluded clients on TB treatment and those specifically seeking free CXR. As for logistical reasons, a maximum of 40 clients could be enrolled per PHC per day, systematic random sampling was applied.

From the COVID-19 vaccine study, we purposely selected healthy controls without any TB symptom who have been followed up for *>* 1 year and showed no signed of any illnesses. From these subjects, we access their archived serum samples which were collected at the beginning of study.

### Study Procedure

Selected serum specimens and medical records were collected. CRP and SAA levels were measured by Fluorescence Immunoassay by Wondfo (Guangzhou Wondfo Biotech Co., Ltd, China), which is a NPOCT fluorescence immunochromato-graphic analyzing system to test biomarkers or other protein using antigen-antibody reaction.(19,20)

IP-10 levels were measured using enzyme-linked immunosorbent assay (ELISA) using LEGEND MAX™ ELISA Kit Human CXCL10 (IP-10) (BioLegend, USA). This kit designed for IP-10 quantitation of human IP-10 from cell culture supernatant, urine, saliva, serum, plasma and other biological fluids and validated with ready to use reagents.(21)

### Statistical Analysis

Baseline characteristics were shown using descriptive statistics. Continuous variables were assessed for normality using the Shapiro–Wilk test and shown as mean ± standard deviation or median (interquartile range), and categorical variables were shown as frequencies and percentages. Comparisons among TB-positive, probable TB, and healthy control groups were analyzed using one-way ANOVA or the Kruskal–Wallis tests, followed by Dunn’s post hoc test. Comparison among groups using independent t-tests or Mann–Whitney U tests and categorical variables was analyzed using Chi-square or Fisher’s tests.

Serum CRP, SAA, and IP-10 levels were compared among diagnostic groups. Receiver operating characteristic (ROC) curve analysis was conducted to evaluate biomarker performance to distinguish in TB-positive subjects from probable TB cases, healthy controls, and the combined non-TB group. The area under the curve (AUC), 95% confidence interval, optimal cut-off value (Youden index), sensitivity, specificity, and diagnostic validity were calculated for each biomarker.

Combined biomarker performance was evaluated using an either-positive approach, in which the test was considered positive if either CRP or SAA exceeded its respective optimal cut-off. Diagnostic validity was calculated as the proportion of subjects correctly classified. Statistical significance was defined as p¡0.05. Analyses were performed using SPSS, GraphPad Prism, and MedCalc.

### Ethical Consideration

This Study was approved by Hasan Sadikin Hospital Ethical Committee, Bandung (approval number: DP.04.03/D.XIV.6.5/21/2026) on January 2026. EVIDENT study was approved by Padjadjaran University Ethical Committee, Bandung (approval number:5/UN6.KEP/EC/2026) on February 2026. COVID-19 vaccine study was approved by Padjadjaran University Ethical Committee, Bandung (approval number 56/UN.6KEP/EC/2023) on September 2023.

## 3. Results

This study enrolled 173 subjects categorized in 3 groups: Tuberculosis group (n=57), Probable tuberculosis group (n=56) and healthy control (n=60). Baseline characteristics including age, sex, symptoms, Body Mass Index (BMI), sputum culture result, Gene Xpert results and chest X-ray findings are shown in Table 1. There is no significant difference regarding the age and gender proportion. There is also no significant difference among the proportion of smoking among the Confirmed TB compared to probable TB. In clinical manifestation, only cough duration and BMI are statistically significant between the groups.

**Table 1.** Baseline Characteristics of Study Subjects.

| **Baseline Characteristics** | **Total Subjects (n=173)** | **Tuberculosis (n=57)** | **Probable TB (n=56)** | **Healthy Control (n=60)** |
|---|---|---|---|---|
| Age (years), median (range) | 43 (18–74) | 39 (18–72) | 44 (18–74) | 43 (19–66) |
| Sex, n (%) |  |  |  |  |
| Male | 106 (61.3) | 38 (66.7) | 31 (55.4) | 37 (61.7) |
| Female | 67 (38.7) | 19 (33.3) | 25 (44.6) | 23 (38.3) |
| Symptoms, n (%) |  |  |  |  |
| Cough | 113 (100.0) | 57 (100.0) | 56 (100.0) | 0 (0) |
| Fever | 77 (68.1) | 40 (70.2) | 37 (66.1) | 0 (0) |
| Night sweats | 70 (61.9) | 38 (66.7) | 32 (57.1) | 0 (0) |
| Weight loss | 78 (69.0) | 45 (78.9) | 33 (58.9)* | 0 (0) |
| Chest pain | 75 (66.4) | 38 (66.7) | 37 (66.1) | 0 (0) |
| Lethargy | 90 (79.6) | 47 (82.5) | 43 (76.8) | 0 (0) |
| Shortness of breath | 83 (73.5) | 43 (75.4) | 40 (71.4) | 0 (0) |
| Hemoptysis | 14 (12.4) | 6 (10.5) | 8 (14.3) | 0 (0) |
| Decreased appetite | 58 (51.3) | 31 (54.4) | 27 (48.2) | 0 (0) |
| Smoking status, n (%) |  |  |  |  |
| Current smoker | 42 (37.2) | 21 (36.8) | 21 (37.5) | nd |
| Former smoker | 13 (11.5) | 8 (14.0) | 5 (8.9) | nd |
| Never smoker | 58 (51.3) | 28 (49.1) | 30 (53.6) | nd |
| Cough duration (days), median (range) | 30 (14–270) | 30 (14–270) | 30 (14–210)* | 0 (0) |
| BMI (kg/m<sup>2</sup>), mean ± SD | 20.28 ± 3.68 | 19.17 ± 3.04 | 21.41 ± 3.95* | 23.7 ± 4.04 |
| Sputum culture (MGIT), n (%) |  |  |  |  |
| Positive | 57 (50.4) | 57 (100.0) | 0 (0.0)* | nd |
| Negative | 56 (49.6) | 0 (0.0) | 56 (100.0)* | nd |
| Molecular rapid test (GenXpert), n (%) |  |  |  |  |
| MTB detected, Rif resistance detected | 1 (0.9) | 1 (1.8) | 0 (0.0)* | – |
| MTB detected, Rif resistance not detected | 53 (46.9) | 53 (93.0) | 0 (0.0)* | – |
| MTB trace detected, Rif resistance indeterminate | 3 (2.7) | 3 (5.3) | 0 (0.0)* | – |
| MTB not detected | 56 (49.6) | 0 (0.0) | 56 (100.0) | – |
| Chest X-ray findings, n (%) |  |  |  |  |
| Suggestive of TB | 76 (43.9) | 57 (100.0) | 19 (33.9) | – |
| Abnormal – non-TB | 17 (15.0) | 0 (0.0) | 17 (30.4) | – |
| Normal | 20 (17.7) | 0 (0.0) | 20 (35.7) | – |

**Notes:** Age and cough duration are presented as median (range) due to non-normal distribution. BMI is presented as mean ± SD because of normal distribution. P-values for numerical variables were analyzed using the Kruskal–Wallis test for age, Mann–Whitney test for cough duration, and independent t-test for BMI. Categorical variables were analyzed using the Chi-square test. \* indicates statistically significant differences (p<0.05). nd: no data.

graphicx booktabs array

We present the numerical distribution of biomarker level across the three diagnostic groups in Table 2. CRP biomarker, a statistically significant difference was observed among the diagnostic groups (p ¡0.001). These results display a marked elevation of CRP levels in subjects with active tuberculosis compared with probable tuberculosis subjects and healthy controls. However, there is no statistically significant in the value of CRP between the probable TB patient and the healthy controls (Figure 1a).

**Table 2.**
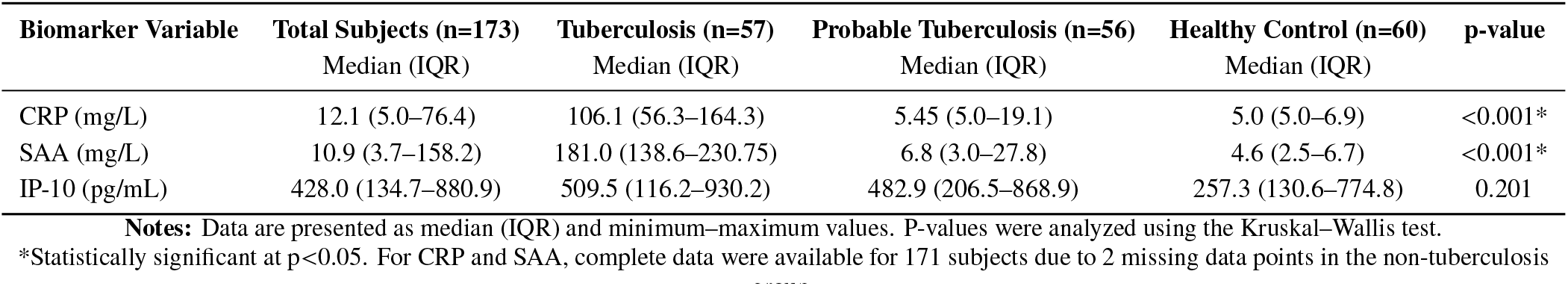
Biomarker Distribution According to Diagnostic Groups.

| **Biomarker Variable** | **Total Subjects (n=173)** | **Tuberculosis (n=57)** | **Probable Tuberculosis (n=56)** | **Healthy Control (n=60)** | **p-value** |
|---|---|---|---|---|---|
|  | Median (IQR) | Median (IQR) | Median (IQR) | Median (IQR) |  |
| CRP (mg/L) | 12.1 (5.0–76.4) | 106.1 (56.3–164.3) | 5.45 (5.0–19.1) | 5.0 (5.0–6.9) | <0.001* |
| SAA (mg/L) | 10.9 (3.7–158.2) | 181.0 (138.6–230.75) | 6.8 (3.0–27.8) | 4.6 (2.5–6.7) | <0.001* |
| IP-10 (pg/mL) | 428.0 (134.7–880.9) | 509.5 (116.2–930.2) | 482.9 (206.5–868.9) | 257.3 (130.6–774.8) | 0.201 |

**Notes:** Data are presented as median (IQR) and minimum–maximum values. P-values were analyzed using the Kruskal–Wallis test.
\*Statistically significant at p<0.05. For CRP and SAA, complete data were available for 171 subjects due to 2 missing data points in the non-tuberculosis [illegible]

**Figure 1a.**
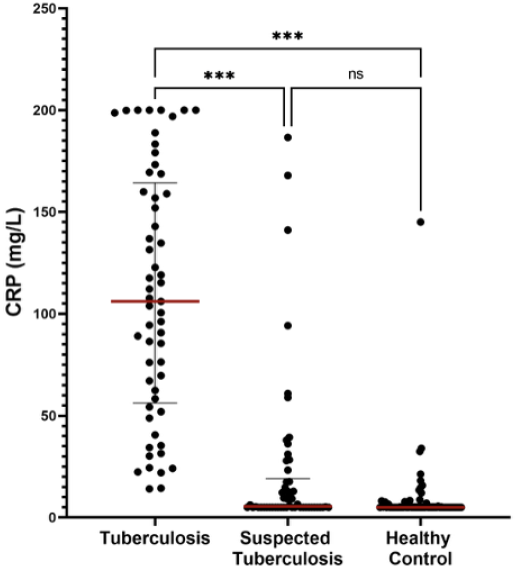
Distribution of serum CRP among the tuberculosis, probable tuberculosis, and healthy control groups.

**Figure 1b.**
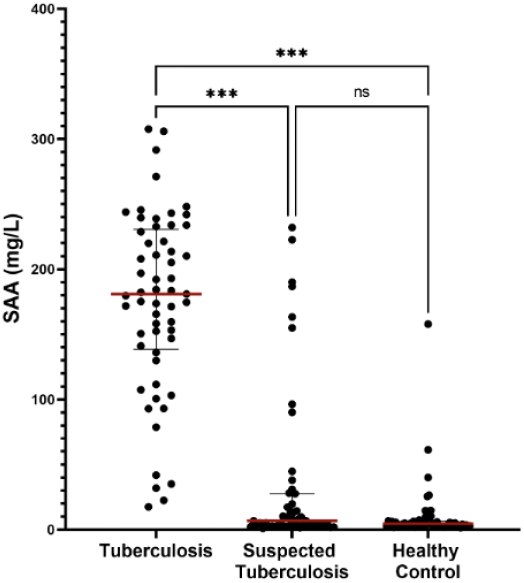
Distribution of serum SAA Levels among the tuberculosis, probable tuberculosis, and healthy control groups.

**Figure 1c.**
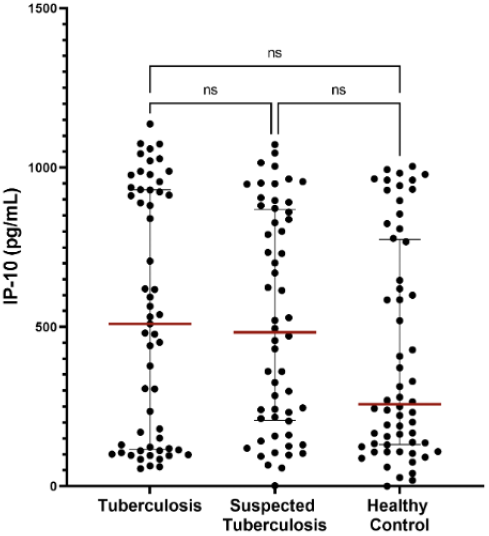
Distribution of serum IP-10 among the tuberculosis, probable tuberculosis, and healthy control groups.

There are significant differences in SAA distribution among the groups (p ¡0.005) (Figure 1b). These differences indicate that SAA levels were also markedly elevated in subjects with active tuberculosis. The increase in SAA in the tuberculosis group appeared seems to be higher than that of CRP, suggesting that this biomarker may possess strong discriminatory potential for differentiating active tuberculosis from non-tuberculosis subjects and healthy controls.

In contrast to CRP and SAA, the IP-10 biomarker did not display a statistically significant difference in distribution among the diagnostic groups (p = 0.201), with a very wide range of values across all groups showing lack of value as diagnostic marker. This condition may explain why no statistically significant difference was identified (Figure 1c), despite the numerically higher median IP-10 level observed in the tuberculosis group.

In figure 2a,2b and 2c we showed that overall, across all three comparative scenarios, SAA consistently presented the highest AUC values to distinguish confirmed TB to probable TB, healthy controls and combination of probable TB and healthy controls (AUC 0.928, 0.991, and 0.961 respectively). With a slightly lower AUC, CRP also provided high AUC at 0.921, 0.982, and 0.953 respectively. In contrast, IP-10 demonstrated AUC values ranging only from 0.508 to 0.577 across all analyses with no statistically significant differences. Table 3 demonstrated specificity and specificity combination of CRP and SAA compared to individual biomarker. In all three group comparison, both CRP and SAA showed sensitivity above 90% and specificity above 70% in distinguishing confirmed TB versus all probable tb, healthy control and combined probable TB and healthy control group. Combination of CRP (cutoffs 21.3 mg/L) and SAA (cutoffs 31.0 mg/L) increases the sensitivity up to 100% while keeping the specificity above 70%.

**Table 3.** ROC Analysis of CRP, SAA, and IP-10 for Differentiating TB-Positive Subjects from Suspected TB, Healthy Controls, and the Combined Non-TB Group.

| **Comparison Group** | **Biomarker** | **Sensitivity (%)** | **Specificity (%)** | **NPV** | **PPV** | **Accuracy (%)** |
|---|---|---|---|---|---|---|
| TB-positive vs suspected TB | CRP >17.7 | 96.5 | 75.9 | 80.9 | 95.3 | 86.5 |
|  | SAA >31.0 | 96.5 | 81.5 | 84.6 | 95.7 | 89.2 |
|  | IP-10 >905.2 | 31.6 | 83.9 | 66.7 | 54.7 | 57.5 |
|  | **CRP >17.7 or SAA >31.0** | **100.0** | **72.2** | **79.2** | **100.0** | **86.5** |
| TB-positive vs healthy controls | CRP >21.3 | 96.5 | 95.0 | 94.8 | 96.6 | 95.7 |
|  | SAA >14.8 | 100.0 | 91.7 | 91.9 | 100.0 | 95.7 |
|  | IP-10 >428.0 | 57.9 | 63.3 | 60.0 | 61.1 | 60.7 |
|  | **CRP >21.3 or SAA >14.8** | **100.0** | **91.7** | **91.9** | **100.0** | **95.7** |
| TB-positive vs suspected TB + healthy controls | CRP >21.3 | 96.5 | 86.0 | 77.5 | 98.0 | 89.5 |
|  | SAA >31.0 | 96.5 | 88.6 | 80.9 | 98.1 | 91.2 |
|  | IP-10 >880.6 | 35.1 | 80.2 | 46.5 | 71.5 | 65.3 |
|  | **CRP >21.3 or SAA >31.0** | **100.0** | **83.3** | **75.0** | **100.0** | **88.9** |

**Notes:** Accuracy was calculated as the proportion of correctly classified subjects among all subjects in each comparison group. Cut-off values represent the optimal thresholds based on the Youden index. For the combined CRP or SAA variable, a positive result was defined as either CRP or SAA being above the specified cut-off.

**Figure 2a.**
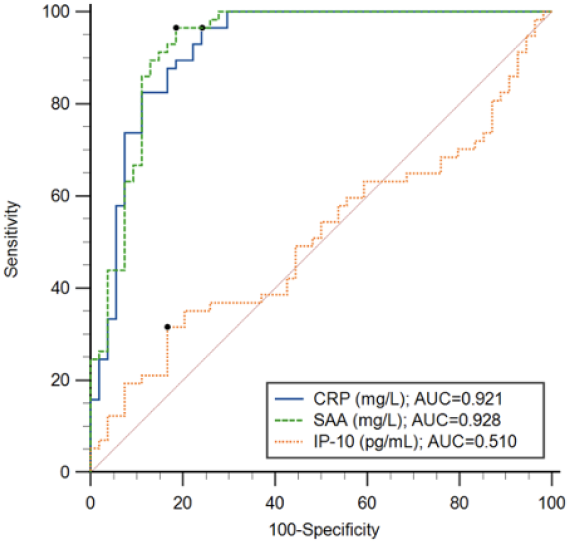
ROC curves of CRP, SAA, and IP-10 for differentiating TB-positive subjects (n=57) from probable TB (n=58) subjects.

**Figure 2b.**
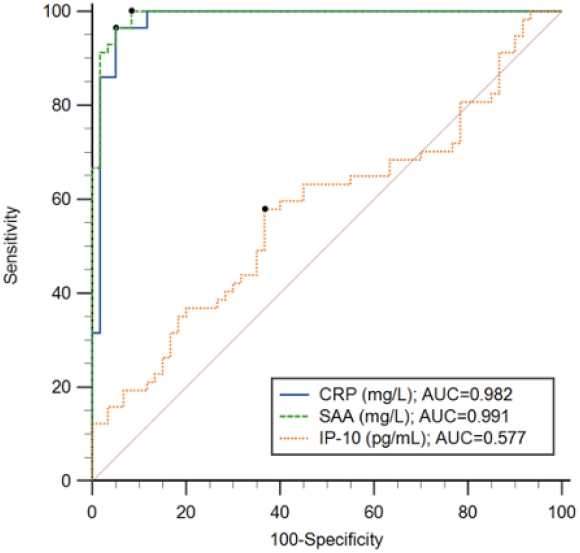
ROC curves of CRP, SAA, and IP-10 for differentiating TB-positive subjects (n=57) from Healthy Control (n=60).

**Figure 2c.**
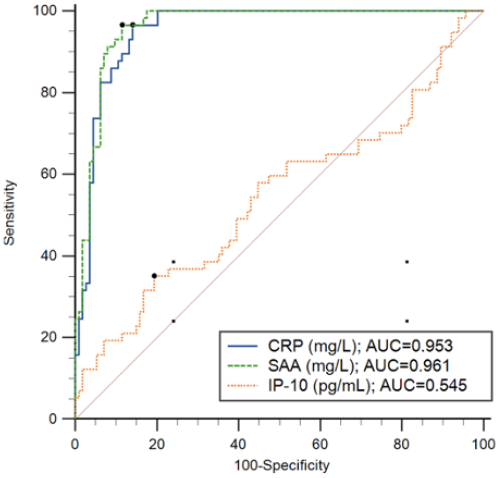
ROC curves of CRP, SAA, and IP-10 for differentiating TB-positive subjects (n =57) from probable TB + Healthy Control (n=116)

## 4. Discussion

Overall, CRP and SAA tests showed excellent sensitivity and specificity for Tuberculosis screening. With this accuracy, both CRP, SAA and the combination of both with the optimal cut off points could achieve WHO target product profile for TB screening or triage test. Looking at this value, it may seem that these two biomarkers may have a diagnostic performance similar to computer aided digital chest x-ray which has a sensitivity and specificity of 90% and 70.3%.(22) The use of CRP in HIV which has been approved by WHO show the sensitivity ranging from 83% to 93%, and specificity ranging from 69-75%.(12) Our study showed that the performance of our studied biomarker can achieve this target.

Our findings are consistent with previous studies. In the meta-analysis by Santos et al., CRP showed 90% sensitivity and 64% sensitivity in high-TB-burden setting.18 Our study showed higher CRP specificity than the large multi-country cohort reported by Crowder et al., in which CRP achieved 49.7% specificity at 90% sensitivity, although differences in study population, setting, cut-off selection, and reference standard may explain part of this difference.(22) For SAA, Jiang et al. reported 69.7% sensitivity and 96.4% specificity for distinguishing patients with TB from healthy controls. However, SAA concentrations were lower in TB than in pneumonia or COPD, indicating that SAA alone is not specific for TB when compared with other inflammatory respiratory diseases.(23). Meanwhile, our IP-10 results were different from previous studies. Santos et al. shows validity studies varies, sensitivity ranging from 78-96%, and specificity 20-91%.

In TB, CRP is a non-specific host-response marker rather than a pathogen-specific diagnostic marker. The severity of TB disease and the mycobacterial burden determined by sputum acid-fast bacilli (AFB) examination have been associated with increased CRP levels and other proinflammatory cytokines.7 Besides the severity of TB, several factors have been reported affecting the degree of CRP increase, some of them include, Mtb strain, ethnicity and site of TB infection.24 Patient with pulmonary, pleural, pericardial, and abdominal TB demonstrating higher CRP concentrations than lymph node, cutaneous, soft tissue, or central nervous system TB.8 In patients with TB, CRP levels decline rapidly within 1–2 weeks after initiation of antituberculosis therapy. However, in some patients, CRP levels do not return to normal within 2 months due to persistent severe inflammation and are associated with failure of sputum culture conversion and the presence of large cavitary lesions on chest radiography, which may be related to paradoxical reactions during antituberculosis treatment.9 In line with these data, CRP concentrations were higher in participants with active TB in our study.

SAA has potential utility as a diagnostic biomarker in infectious diseases, including bloodstream infection, sepsis, pneumonia, tuberculosis, and COVID-19; inflammatory disorders such as rheumatoid arthritis, lupus, and inflammatory bowel disease; cardiovascular diseases; malignancies; amyloidosis; and metabolic disorders. 25,14 SAA has not been extensively investigated, and previous studies have been limited, including research conducted by Jiang et al. in China, in which TB patients were also compared with patients with pneumonia.11 Several studies have demonstrated that the magnitude of the SAA response is greater than that of CRP, suggesting that SAA may be more sensitive and potentially more useful than CRP as a biomarker of acute inflammatory responses.12 Both CRP and SAA tested in this validation study are near point-of-care tests compatible with serum, plasma or whole blood sample from phlebotomy as well as finger-prick access. We believe it can be highly practical for mass community screening or immediate outpatient deployment. Notably, both assays feature a rapid turnaround time of only three minutes per test, which substantially reduces patient waiting times in primary care triage settings. Furthermore, the complete assay kits can be safely stored at room temperature, minimizing infrastructural and cold-chain requirements for decentralized diagnostic points

IP-10 is a plausible host-response biomarker for TB, where control of Mtb depends partly on coordinated Th1 immunity. IP-10 may have potential utility in differentiating latent TB from active TB; however, IP-10 reflects IFN--driven inflammation rather than TB-specific pathology, and increased concentrations may occur in both active TB and latent infection, and thus, further studies are still required.14 A study conducted in Indonesia demonstrated lower IFN-levels in patients with TB disease compared with findings from other studies. This discrepancy has led to the hypothesis that reduced IFN-expression may contribute to the progression of TB infection into active disease with clinical manifestations.16 Lower IFN-levels may therefore be associated with the relatively low IP-10 levels observed in the present study.

This study has several limitations. The sample size was relatively small and participants were purposively selected, which may limit generalizability. Although the study population reflects TB cases commonly encountered in community settings, diagnostic performance may differ in broader populations. Healthy controls were defined by the absence of TB symptoms without confirmation by chest radiography, molecular testing, or culture. In addition, individuals with other infectious or inflammatory diseases and immunocompromising conditions, including HIV infection, diabetes mellitus, or immunomodulatory treatment, were excluded. Therefore, the diagnostic performance of these biomarkers in populations with concurrent inflammation or immune dysfunction requires further investigation.

## 5. Conclusions

In conclusion, our finding suggest that SAA and CRP biomarkers, but not IP-10, either individually or in combination, may be used as screening tests for tuberculosis that meet the World Health Organization (WHO) target product profile. Among the single biomarkers, SAA demonstrated the best overall sensitivity and specificity, while the most effective combination was CRP and SAA. We found no adequate diagnostic value of IP-10 in our study. Future studies should examine the diagnostic performance of CRP and SAA in comparison with chest Xray as well as other screening tools. Larger trial with a whole spectrum of TB disease in a community setting should also be conducted to understand their real implementation impacts.

## Data Availability

All data produced in the present study are available upon reasonable request to the authors.

## Author Contribution

NI: Conceptualization, Methodology, Investigation, Data curation, Formal analysis, Validation, Writing – original draft, Writing – review editing, Project administration; TPS: Methodology, Data curation, Validation, Writing – review editing; MK: Methodology, Investigation, Data curation, Validation, Writing – review editing; RCK: Methodology, Investigation, Data curation, Validation, Writing – review editing; IDK: Methodology, Investigation, Validation, Writing – review editing; DKT: Methodology, Investigation, Validation, Writing – review editing; BA: Conceptualization, Validation, Writing – review editing, Supervision, Project administration.

## Acknowledgements

We would like to thank all the participants of EVIDENT and LIVZON. We also would like to thank Sri Hartati and Chrisan Bimo for assisting in subject selection of EVIDENT and LIV-ZON, Adnes Mareta and Keyvino Rettob for assisting in CRP and SAA measurement. The EVIDENT study was supported by Gates Foundation.

